# The effects of polygenic risk for psychiatric disorders and smoking behaviour on psychotic experiences in UK Biobank

**DOI:** 10.1101/2019.12.19.19015339

**Authors:** Judit García-González, Julia Ramírez, David M. Howard, Caroline H Brennan, Patricia B. Munroe, Robert Keers

**Author notes:** Equally contributed (Patricia B. Munroe, Robert Keers). Address correspondence to Judit García-González, School of Biological and Chemical Sciences, Queen Mary, University of London, London E1 4NS, UK.

## Abstract

While psychotic experiences are core symptoms of mental health disorders like schizophrenia, they are also reported by 5-10% of the population. Both smoking behaviour and genetic risk for psychiatric disorders have been associated with psychotic experiences, but the interplay between these factors remains poorly understood. We tested whether smoking status, maternal smoking around birth, and number of packs smoked/year were associated with lifetime occurrence of three psychotic experiences phenotypes: delusions (n=2 067), hallucinations (n=6 689), and any psychotic experience (delusions or hallucinations; n=7 803) in 157 366 UK Biobank participants. We next calculated polygenic risk scores for schizophrenia (PRS_SCZ_), bipolar disorder (PRS_BP_), major depression (PRS_DEP_) and attention deficit hyperactivity disorder (PRS_ADHD_) in 144 818 UK Biobank participants of European ancestry to assess whether association between smoking and psychotic experiences was attenuated after adjustment of diagnosis of psychiatric disorders and the PRSs. Finally, we investigated whether smoking exacerbates the effects of genetic predisposition on the psychotic phenotypes in gene-environment interaction models. Smoking status, maternal smoking, and number of packs smoked/year were associated with psychotic experiences (*p*<1.77×10^-5^). Except for packs smoked/year, effects were attenuated but remained significant after adjustment for diagnosis of psychiatric disorders and PRSs (*p*<1.99×10^-3^). Gene-environment interaction models showed the effects of PRS_DEP_ and PRS_ADHD_ (but not PRS_SCZ_ or PRS_BP_) on delusions (but not hallucinations) were significantly greater in current smokers compared to never smokers (*p*<0.002). There were no significant gene-environment interactions for maternal smoking nor for number of packs smoked/year. Our results suggest that both genetic risk of psychiatric disorders and smoking status may have independent and synergistic effects on specific types of psychotic experiences.

## Introduction

Psychotic experiences, such as hallucinations (unreal visual or auditory perceptions) and delusions (unreal beliefs or impressions, i.e. conspiracy against self, unreal communications or signs), are reported by 5-10% of the general population^1^. Psychotic experiences are core symptoms of severe mental disorders, such as schizophrenia, and therefore may be an indicator of risk for mental health problems even in apparently healthy individuals. Understanding their aetiology could help identify individual susceptibility for different mental health outcomes and develop tailored prevention and intervention strategies.

Genetic factors influence psychotic experiences, explaining between 30-50% of the variance in twin studies ^2,3^ and common genetic variation explaining between 3-17% of the variance in molecular genetic studies ^4,5^. Twin studies also suggest that the genetic component of psychotic experiences is shared with psychiatric disorders such as schizophrenia, bipolar disorder, depression and attention deficit hyperactivity disorder (ADHD)^6^. Large genome-wide studies of psychotic experiences in UK Biobank (UKB) confirm these findings: polygenic risk scores (PRS) of schizophrenia, bipolar disorder, depression, and ADHD explain 0.11%, 0.06%, 0.32%, and 0.05% of the variance in psychotic experiences, respectively ^7^. Linkage disequilibrium score regression (LDSC) indicates the genetic correlation estimates between psychotic experiences and schizophrenia, bipolar disorder, depression and ADHD are 0.21, 0.15, 0.46 and 0.24^7^.

In addition to genetic influences, environmental factors explain between 50-70% of the variance in psychotic experiences ^3^. Smoking is one of the best established environmental risk factors associated with the occurrence of psychotic experiences in case-control studies of schizophrenia and bipolar disorder, as well as in population-based samples. Longitudinal studies suggest that continued smoking is a causal factor for psychotic experiences among people with first episode of psychosis ^8^, schizophrenia, and bipolar disorder patients ^9^. The observation of a dose-response effect in prospective studies ^10-13^, together with recent Mendelian randomization studies ^14^, further support a causal effect of smoking on the occurrence of psychotic experiences. In addition, the associations between lifetime tobacco use and subsequent psychotic experiences ^15,16^ in the general population remained significant after adjusting for other psychiatric disorders ^16^, suggesting that smoking may lead to psychotic experiences independently from mental health status.

Associations between other smoking phenotypes and psychotic experiences are less well-established. There is much less evidence for an increased risk of psychotic experiences in former smokers, leading some to suggest that the effects of smoking are reversible ^17,18^. However, it is still unclear whether these findings are false positives resulting from a lack of power in relatively small samples. Maternal smoking during pregnancy is associated with psychotic experiences in offspring ^19^. However within-family studies suggest that once familial factors are taken into account, maternal smoking during pregnancy has no effect on the risk of psychotic experiences^20^.

Despite evidence showing associations between genetic risk for psychiatric disorders and psychotic experiences ^6,7^, and a number of studies showing associations between smoking and psychotic experiences ^17,21-23^, the interplay between smoking and genetic risk on psychotic experiences remains unknown. There are studies showing that PRSs for psychiatric disorders are associated with smoking ^24-27^. This suggests that smoking may not be only a casual factor in the development of psychotic experiences, but that common genetic factors may affect both smoking and psychotic experiences. However, no studies have tested whether PRSs for psychiatric disorders and smoking make independent contributions to psychotic experiences. Significant gene-environment interactions (GxE) have been reported for several psychiatric traits including schizophrenia and depression ^28-30^, but studies are yet to explore whether there is an interaction between smoking and PRSs for psychiatric disorders on psychotic experiences. That is, whether the effects of genetic risk are greater for individuals exposed to smoking.

Our first objective was to investigate whether smoking status (current, former or never smoker), number of packs smoked per year, and maternal smoking around birth were associated with lifetime delusions or hallucinations or any psychotic experience (delusions or hallucinations) in the UKB cohort. We also tested whether the effects of smoking remained significant after adjustment for diagnosis of depression, ADHD and psychotic disorders (schizophrenia, mania, hypomania, bipolar or manic depression, and schizotypal or delusional disorders), as well as genetic risk for these disorders, captured by PRSs for depression (PRS_DEP_), ADHD (PRS_ADHD_), schizophrenia (PRS_SCZ_) and bipolar disorder (PRS_BP_). Our second objective was to investigate whether the effects of smoking on psychotic experiences were exacerbated by genetic risk by testing additive and multiplicative GxE between these variables. We interrogated whether being a current, former or never smoker; smoking more packs of cigarettes per year; or being exposed to maternal smoking around birth influenced the effects of PRSs on having hallucinations, delusions, or any of the two. When significant interactions between smoking and PRS were identified, we carried out sensitivity analyses excluding people with psychotic disorders. An overview of the study design is provided in Figure 1.

**Figure 1.**
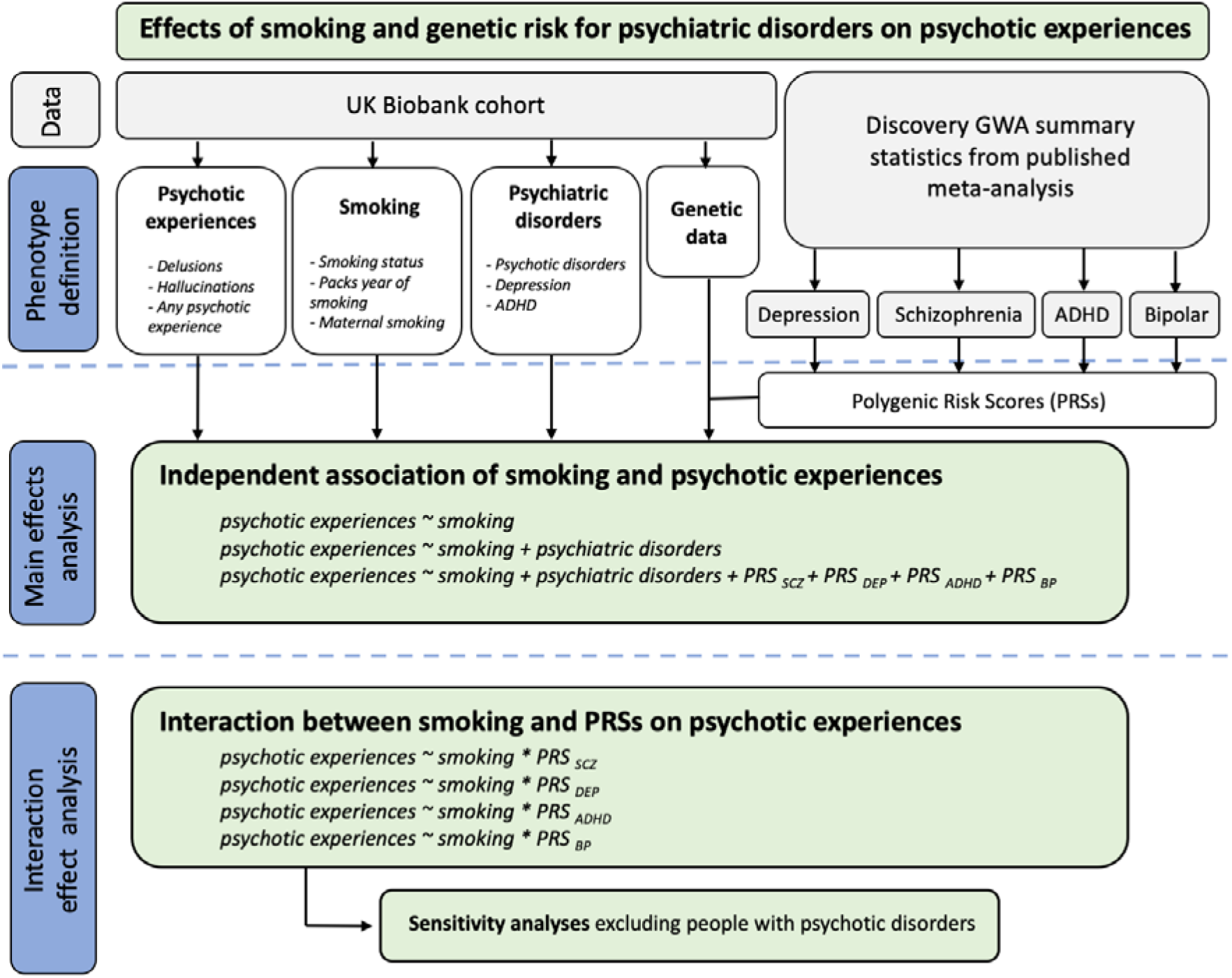
Detailed flowchart of the analytical approach. UKB: UK Biobank cohort. PRSs: polygenic risk scores for schizophrenia (PRS_SCZ_), depression (PRS_DEP_), ADHD (PRS_ADHD_) and Bipolar disorder (PRS_BP_). GWA: genome wide association. ADHD: Attention deficit hyperactivity disorder.

## Methods

### UK Biobank (UKB) sample

The sample was drawn from UKB, a population-based cohort from the United Kingdom with ~500 000 participants. From those, 157 366 participants completed an online mental health questionnaire. A detailed description of the mental health questionnaire has been provided elsewhere ^31^. Ethical approval was provided by UKB (Application ID#42423) and all participants gave written consent.

### Measures

#### Psychotic experiences

Psychotic experiences were categorised based on the Composite International Diagnostic Interview included in the mental health questionnaire. Three outcomes were defined: *delusions, hallucinations*, and *any psychotic experience*. Delusions were ascertained as individuals reporting lifetime delusions of reference and/or persecutory delusions (Field IDs: 20474, 20468). Hallucinations were ascertained as individuals reporting lifetime visual and/or auditory hallucinations (Field IDs: 20471, 20463). Any lifetime psychotic experiences were ascertained as a positive response to any of the four symptom questions (Field IDs: 20474, 20468, 20471, 20463). Controls were defined as individuals that did not endorse lifetime psychotic experiences. A more detailed description of the outcome variables is provided in the Supplementary Tables 1-3.

#### Smoking

Three smoking outcomes were defined using the UKB baseline questionnaire: smoking status (Field ID 20116), pack years of smoking (Field ID 20161) and maternal smoking around birth (Field ID 1787). The smoking status phenotype consisted of *never smoker, former smokers* and *current smokers*. Pack years of smoking was calculated for former and current smokers and it was defined as number of cigarettes smoked per day, divided by twenty, multiplied by the number of years of smoking. Cases for maternal smoking were defined as individuals endorsing “Yes” for the question *“Did your mother smoke regularly around the time when you were born?”*.

#### Other psychiatric disorders

Lifetime depression, ADHD, schizophrenia, mania, hypomania, bipolar or manic depression, and any other type of psychotic disorder were defined using both the UKB baseline and mental health questionnaires (Supplementary Tables 4 and 5).

#### UKB genetic dataset

The data release contained 488 377 individuals genotyped on either the UKB Axiom or the UK BiLEVE Axiom arrays. Genotype data were imputed centrally by UKB with IMPUTE2 using the Haplotype Reference Consortium panel^32^. UKB also provided metrics for quality control that were used to exclude individuals with poor genotype call rate (>5%) and discordance between the self-reported sex and the sex inferred from the genotypes. The first two principal components provided by UKB were used in a k-means clustering algorithm ^33^ to identify a genetically homogeneous subsample. Related individuals were identified for each phenotype separately using the R package ‘ukbtools’ ^34^ and one individual from related pairs/trios were excluded (kinship coefficient > 0.0884). Details on the quality control (QC) of genetic data and sample sizes after QC exclusion are provided in Supplementary Table 6.

#### PRSs

We calculated PRS_SCZ_, PRS_DEP_, PRS_ADHD_ and PRS_BP_ for each UKB participant using the software PRSice-2 ^35^ and summary statistics from four recent genome-wide association studies GWAS of these disorders (Supplementary material) ^36-39^. Logistic regression models were used to explore the association between PRS_SCZ_, PRS_DEP_, PRS_ADHD_ and PRS_BP_ and psychotic experiences using PRSice-2 ^35^. Analyses included the first ten principal components and genotyping batch as covariates.

#### Main effect analyses

Regression models were constructed to assess the association between maternal smoking, smoking status and number of packs smoked per year and psychotic experiences. To test whether the association between smoking and psychotic experiences was independent of diagnosis for psychiatric disorders and genetic predisposition, models were rerun adjusting for: diagnosis for psychotic disorders, depression, and ADHD, as well as the PRS with highest predictive ability (p-value threshold = 0.2) on psychotic experiences.

#### Interaction effect analyses

We explored whether there were GxE between each of the PRSs and smoking on psychotic experiences (i.e. whether the effects of genetic risk differed among current, former or never smokers, among smokers that consumed more packs of cigarettes per year, and among individuals whose mothers smoked around time of participants’ birth). Interactions were tested on the multiplicative and additive scales using logistic and linear regressions, respectively. Interactions on the *multiplicative scale* assess whether the joint effect of the PRS and smoking is greater than the *product* of their individual effects. Interactions on the *additive scale* assess whether the joint effect of smoking and the PRS is greater than the *sum* of their individual effects. Modelling multiplicative and additive GxE using linear and logistic regressions has been described elsewhere^29,30,40^.

In the GxE regression models, we included individuals from UKB with genetic, smoking and psychotic experiences data (n = 143 320 for any psychotic experience, n = 143 043 for hallucinations, and n = 143 245 for delusions). Independent variables were the PRS_DEP_, PRS_ADHD_, PRS_SCZ_ or PRS_BP_ for each individual, smoking phenotype, ten principal components and genotyping batch. Dependent variables were lifetime occurrence of hallucinations, delusions or any psychotic experience. Models also included the interaction of smoking and PRS with genotyping batch and the principal components, to control for the effect of these covariates on the interaction term ^41^. To examine whether the GxE was significant after multiple testing correction (*p* < 0.0021 based on 24 models under additive and multiplicative models: 0.05/24 = 0.0021), we used the anova function from R package “car”. All models were constructed in R (3.5.1).

#### Sensitivity analyses

The inclusion of individuals with psychotic disorders (Supplementary Table 4) may inflate the variance explained by the PRSs and lead to false positive GxE. Therefore, we repeated the significant interaction effect analysis after excluding individuals that met criteria for psychotic disorders.

### Code availability

Analytical code used to define outcomes and regression models for this project is available at https://github.com/iuditperala/PRSxSmoking.

## RESULTS

157 366 individuals completed the mental health questionnaire. From those, 2 067 reported delusions, 6 689 hallucinations and 7 803 any psychotic experience. 953 individuals had both lifetime delusions and hallucinations. Sample sizes for controls were 155 182, 150 336 and 149 289 for delusions, hallucinations, and any psychotic experience, respectively.

For the smoking variables derived from the full cohort, 273 542 individuals reported that they never smoked, 173 072 were former smokers and 52 979 were current smokers. The mean number of packs per year smoked was 23.4 (SD=18.8). 126 632 individuals reported that their mothers had smoked around the participant’s birth, whereas 306 266 reported the opposite. Table 1 includes demographic, psychiatric and behavioural characteristics of the sample *before* genetic data QC. Supplementary Table 7 shows the number of individuals with psychotic experiences stratified by smoking status, maternal smoking and mean number of packs smoked per year *after* genetic data QC.

**Table 1.**
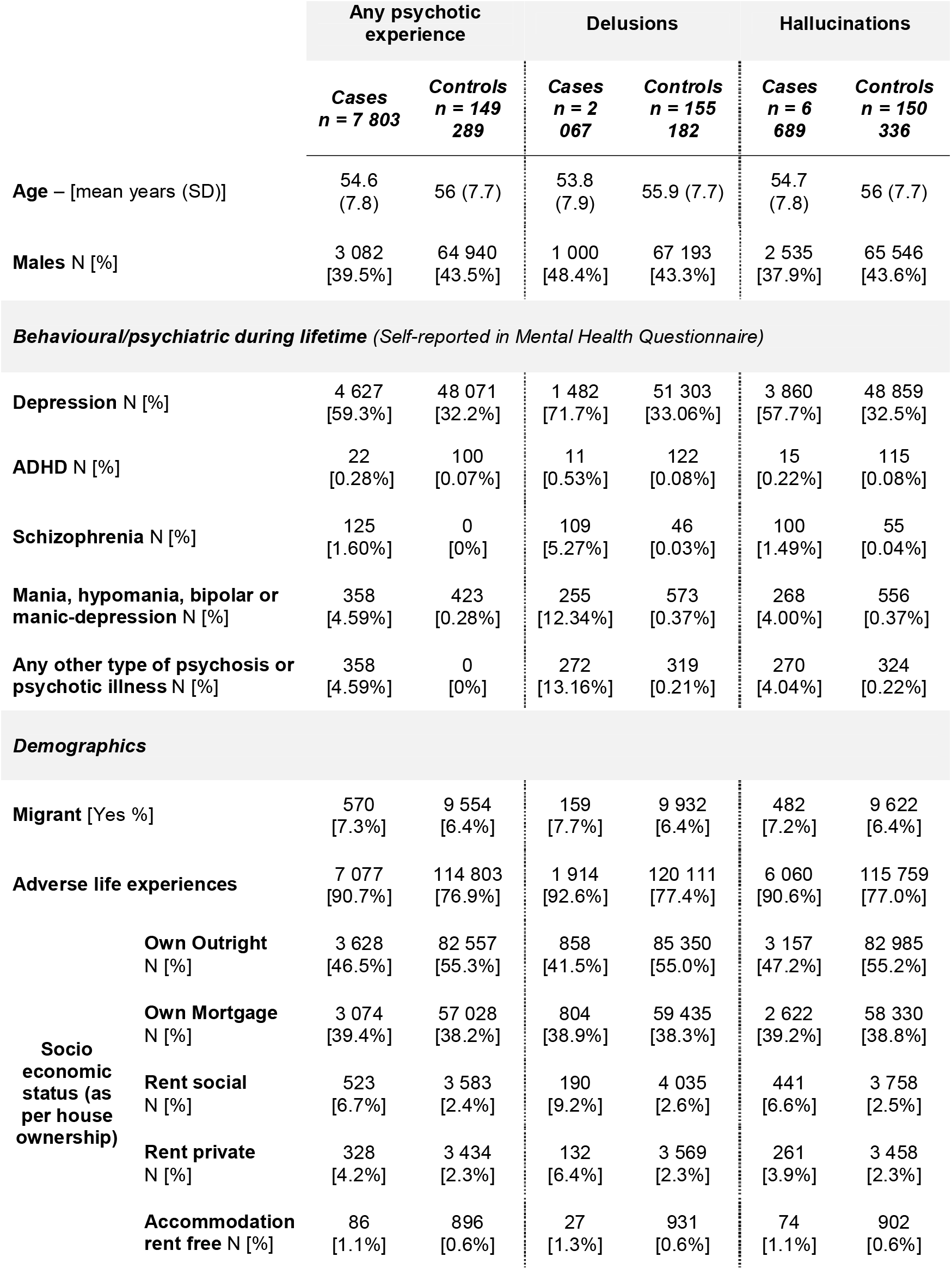
Demographic, psychiatric and behavioural characteristics of the sample.

### Smoking and psychotic experiences

The odds of having delusions, hallucinations or any psychotic experience was significantly higher in current or former tobacco users when compared with non-users, and in participants whose mothers smoked around the time of birth (Table 2). There was a positive linear relationship between the number of packs of cigarettes and risk of psychotic experiences (Table 2). Associations between smoking and the likelihood of having psychotic experiences were strongest for continued tobacco smoking (current smoking vs never smoked). Associations were attenuated after adjustment for diagnosis of psychotic experiences, depression, and ADHD but remained significant. Only packs smoked per year on occurrence of delusions did not pass the significance threshold after multiple testing correction (*p* < 0.0125) (Table 2).

**Table 2.** Phenotypic relationships between smoking and psychotic experiences within UKB. Logistic regression models were used to assess the association between smoking and three psychotic experiences phenotypes. OR; Odds ratio, *p;* p-value. Significance was declared at *p* < 0.0125 after Bonferroni correction (based on four tests, one per smoking phenotype).

|  | Unadjusted<sup>a</sup> |  | Adjusted for psychiatric disorders<sup>b</sup> |  | Adjusted for psychiatric disorders and PRSs<sup>c</sup> |  |
| --- | --- | --- | --- | --- | --- | --- |
|  | OR (95% CI) | *p* | OR (95% CI) | *p* | OR (95% CI) | *p* |
| ***Any psychotic experience*** |  |  |  |  |  |  |
| Former vs never smoking | 1.22 (1.16-1.28) | $4.99 \times 10^{-14}$ | 1.16 (1.10-1.22) | $2.89 \times 10^{-8}$ | 1.15 (1.09-1.21) | $2.87 \times 10^{-7}$ |
| Current vs never smoking | 1.76 (1.62-1.91) | $4.76 \times 10^{-42}$ | 1.47 (1.35-1.60) | $3.76 \times 10^{-19}$ | 1.45 (1.33-1.58) | $1.08 \times 10^{-17}$ |
| Maternal smoking | 1.26 (1.19-1.33) | $5.89 \times 10^{-17}$ | 1.20 (1.14-1.27) | $5.87 \times 10^{-11}$ | 1.19 (1.13-1.26) | $6.98 \times 10^{-10}$ |
| Packs smoked per year | 1.006 (1.004-1.008) | $4.61 \times 10^{-7}$ | 1.004 (1.001-1.006) | $2.95 \times 10^{-3}$ | 1.003 (1.001-1.006) | $7.17 \times 10^{-3}$ |
| ***Delusions*** |  |  |  |  |  |  |
| Former vs never smoking | 1.28 (1.16-1.41) | $1.68 \times 10^{-6}$ | 1.18 (1.07-1.31) | $1.29 \times 10^{-3}$ | 1.17 (1.05-1.29) | $3.90 \times 10^{-3}$ |
| Current vs never smoking | 2.21 (1.9-2.55) | $9.74 \times 10^{-27}$ | 1.61 (1.38-1.87) | $9.41 \times 10^{-10}$ | 1.57 (1.35-1.83) | $8.13 \times 10^{-9}$ |
| Maternal smoking | 1.28 (1.15-1.42) | $3.17 \times 10^{-6}$ | 1.20 (1.08-1.34) | $8.09 \times 10^{-4}$ | 1.19 (1.06-1.32) | $2.06 \times 10^{-3}$ |
| Packs smoked per year | 1.009 (1.005-1.013) | $4.73 \times 10^{-6}$ | 1.005 (1.000-1.009) | 0.0383 | 1.004 (0.999-1.008) | 0.0740 |
| ***Hallucinations*** |  |  |  |  |  |  |
| Former vs never smoking | 1.21 (1.14-1.28) | $2.66 \times 10^{-11}$ | 1.15 (1.09-1.22) | $8.05 \times 10^{-7}$ | 1.14 (1.08-1.21) | $4.76 \times 10^{-6}$ |
| Current vs never smoking | 1.76 (1.61-1.92) | $8.08 \times 10^{-37}$ | 1.49 (1.36-1.63) | $2.36 \times 10^{-18}$ | 1.47 (1.34-1.61) | $4.86 \times 10^{-17}$ |
| Maternal smoking | 1.26 (1.19-1.34) | $4.47 \times 10^{-15}$ | 1.21 (1.14-1.28) | $4.66 \times 10^{-10}$ | 1.20 (1.13-1.27) | $3.92 \times 10^{-9}$ |
| Packs smoked per year | 1.005 (1.003-1.008) | $1.77 \times 10^{-5}$ | 1.003 (1.001-1.006) | 0.0101 | 1.003 (1.001-1.006) | 0.0185 |

Note: For the predictor 'packs smoked per year', OR represents the increase in odds ratio *for one unit increase* in the number of packs smoked per year. Unadjusted model <sup>a</sup> assessed the phenotypic relationship between psychotic experiences and smoking using logistic regression models. Adjusted model (b) included as covariates: diagnosis for psychotic disorders, ADHD and depression. Adjusted model (c) included as covariates: diagnosis for psychotic disorders, depression and ADHD, as well as PRS for schizophrenia, PRS for depression, PRS for ADHD, and PRS for Bipolar disorder.

The PRS_SCZ_ - PRS_DEP_, PRS_ADHD_ and PRS_BP_ were significantly associated with psychotic experiences (Figure 2) and with smoking behaviour (Supplementary Figure 1). The proportion of variance in psychotic experiences explained by PRSs was small, the highest was for PRS_SCZ_, which predicted 0.89% of the variance for lifetime delusions (Figure 2A). Results were dependent on the type of psychotic experience: PRS_SCZ_ and PRS_BP_ explained almost double the percentage of the variance for delusions than for hallucinations (Figure 2A and 2D), and p-values for PRS_ADHD_ passed multiple testing correction for hallucinations and any psychotic experience, but not for delusions (Figure 2C).

**Figure 2.**
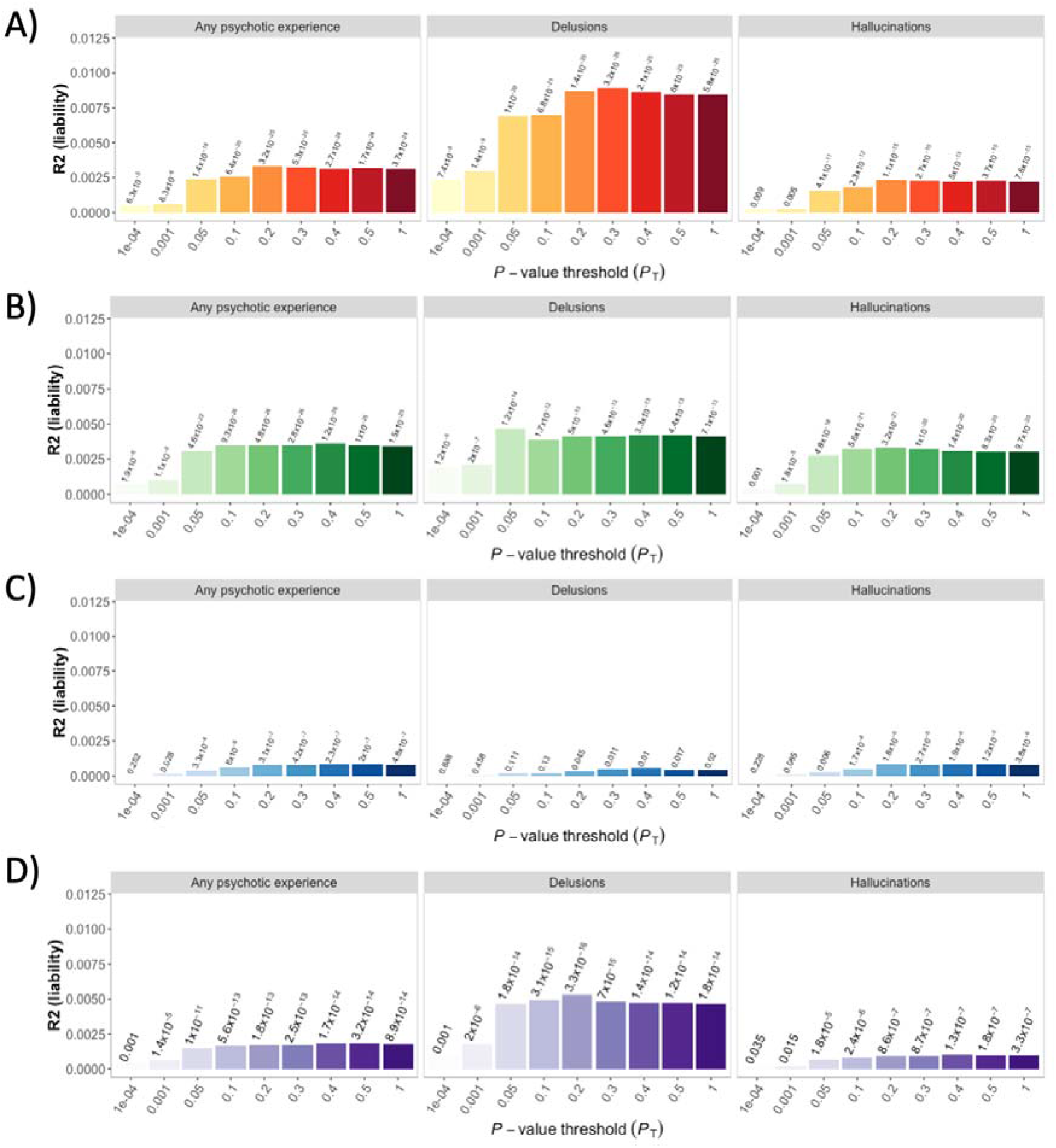
Prediction of psychotic experiences in UKB using PRSs for schizophrenia (Panel A), depression (Panel B), ADHD (Panel C) and bipolar disorder (Panel D). Each panel is divided into types of psychotic experiences. The Y axis represents variance of psychotic experience explained by the PRS in the liability scale (Nagelkerke R^2^). The X axis represents different genetic association p-value thresholds used to build the PRS. The p-values of the association between the PRSs and psychotic experiences are listed above of the bars.

Therefore, it is possible that genetic risk confounds the relationship between smoking and psychotic experiences. To test this hypothesis, we included PRS_SCZ_, PRS_DEP_, PRS_ADHD_ and PRS_BP_ as covariates in the analyses. All the regression models (except packs smoked per year on delusions and hallucinations) remained significant after the inclusion of the PRS_SCZ_, PRS_DEP_, PRS_ADHD_, and PRS_BP_ (Table 2).

### Interaction between genetic risk of psychiatric disorders and smoking on psychotic experiences

We found significant additive interactions between PRS_ADHD_ and PRS_DEP_ (but not PRS_SCZ_ and PRS_BP_) and smoking status (current i/s never smoker) on reporting lifetime delusions. That is, the combined effect of PRS_ADHD_ and smoking, or PRS_DEP_ and smoking was significantly greater than the sum of their individual effects (Figure 3 and Supplementary Table 8 for OR and P values). However, there were no significant interactions between PRSs and smoking status for hallucinations or any psychotic experiences (Figure 3).

**Figure 3.**
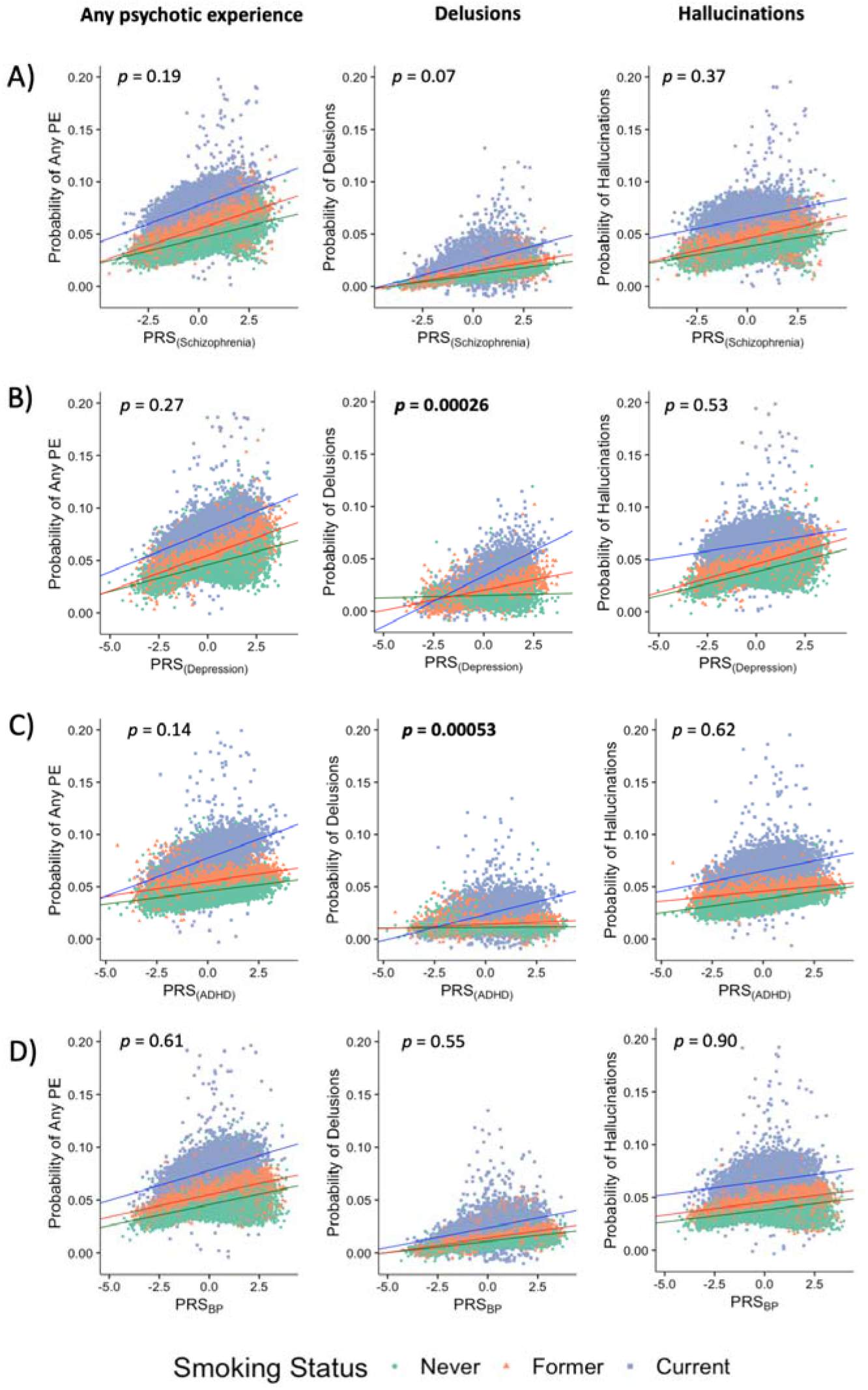
Associations between polygenic risk score for A) schizophrenia (PRS_SCZ_), B) depression (PRS_DEP_), C) ADHD (PRS_ADHD_) and D) bipolar disorder (PRS_BP_), and occurrence of psychotic experiences (PE) under the additive model. Current smokers are shown as blue squares, former smokers as orange triangles, and never smokers as green circles. P values for the PRS x smoking status interactions are shown on the upper part of each plot. Significant associations (*p* < 0.0021 based on 24 tests; 12 additive interactions and 12 multiplicative interactions) are highlighted in bold. The effects of PRS_DEP_ and PRS_ADHD_ on the probability of having delusions are greater among current smokers than former and never smokers.

To ensure that the significant interaction for PRS_ADHD_ and PRS_DEP_ was not entirely driven by participants with a diagnosis of bipolar disorder or schizophrenia, we excluded individuals meeting criteria for diagnosis of psychotic disorders (Supplementary Table 9) and repeated the GxE analyses. All the interaction terms remained significant (Supplementary Figure 2 and Supplementary Table 10).

There were no significant interactions between the PRSs and maternal smoking, nor between the PRSs and number of packs per year (Supplementary Tables 11 and 12).

## Discussion

Our study investigated whether smoking is associated with the occurrence of psychotic experiences independently from diagnosis of psychotic disorders, depression or ADHD, or genetic predisposition for these disorders. We also tested whether the effects of smoking were modulated by the genetic risk in gene-environment interaction models. We found smoking status, maternal smoking and number of packs smoked per year were all significantly associated with increased risk of psychotic experiences. GxE modelling indicated the effects of two PRSs_(dep&adhd)_ on delusions was significantly greater in current smokers compared to never smokers. These results suggest both genetic risk and smoking behaviour contribute to specific types of psychotic experiences.

A few previous studies have assessed PRS-environment interactions on psychotic experiences finding that higher genetic risk exacerbates environmental effects. However, these studies have focused exclusively on genetic risk of schizophrenia and environmental factors such as stress ^42^ and birth weight ^43^. Our study is the first one to explore PRS by environment interactions with smoking on psychotic experiences and the first to explore these effects in the context of genetic risk of depression, bipolar disorder and ADHD. Further, our study had greater power than previous ones because we used at least 30 times larger sample sizes, we calculated multiple PRSs at higher p-values that increased the predictive power of the genetic score, and we used the largest GWAS summary statistics available to date.

The interaction of smoking status with PRS_DEP_, but not with PRS_SCZ_ or PRS_BP_, on predicting delusions is of particular interest. Delusions of reference and persecution are classified as some of the core symptoms of psychotic disorders (i.e. schizophrenia and bipolar disorder) but they are rarely part of the diagnostic criteria for depression^44^. Wooton et al^14^ reported that genetic risk for depression increased smoking, but there was unclear evidence for genetic risk for schizophrenia increasing smoking. Our finding that PRS_DEP_ (but not PRS_SCZ_ or PRS_BP_) interacts with smoking on predicting psychotic experiences points at a stronger relationship between smoking and depression than between smoking and schizophrenia or between smoking and bipolar disorder. Our sensitivity analyses, where the interaction remained significant after excluding people with psychotic disorders, supports this hypothesis.

In line with previous studies, we found significant associations between the smoking phenotypes and psychotic experiences ^10-13,17^_;_ and between PRS_SCZ_ - PRS_DEP_, PRS_ADHD_ and PRS_BP_ and the smoking phenotypes and psychotic experiences ^25,36-45,46^. These findings suggest that the relationship between the smoking variables and psychotic experiences may be confounded by shared genetic influences. However, the effects of smoking remained very similar after adjusting for PRSs, providing little evidence of genetic confounding by common genetic variation associated with schizophrenia, depression, ADHD and bipolar disorder. Previous studies have shown that the effects of smoking on psychotic experiences remain significant after adjustment for psychiatric disorders ^16^. However, this is the first study to show these effects also remain significant after adjustment for genetic risk of specific psychiatric disorders.

The effects of maternal smoking after adjusting for the PRSs are also interesting. Previous within-family studies suggest that, once unmeasured familial factors are taken into account, maternal smoking has little effect on psychotic disorders^20^. In the current study, the effects of maternal smoking persisted following adjustment for PRS_SCZ_ - PRS_DEP_ PRS_ADHD_ and PRS_BP_. This tentatively suggests that genetic risk for these disorders does not confound the relationship between maternal smoking and psychotic experiences. Nevertheless, the PRSs tested only capture a small fraction of the genetic risk for psychiatric disorders. Replication of these effects using more powerful estimates of genetic risk will be required to test this hypothesis.

There are some limitations of this study: it is retrospective meaning it is subject to recalling biases, which occur when participants do not accurately remember past experiences, and this can lead to spurious results. We have relied on self-reported data, which might be affected by different conceptions of what it is a psychotic experience. There were 351 individuals that completed the UKB mental health questionnaire that may have also participated in the Psychiatric Genomic Consortium cohorts analysed by Wray an colleagues^37^. The sample overlap was minimal, with 0.079% for the discovery sample (351/446 238), and 0.242% for the target sample (351/144 818). The overlap was identified using a checksum-based approach as described in Howard et al^47^ and the impact over this overlap is unknown. We reported an interaction between genetic risk and smoking, but the percentage of variance explained by the PRSs was very low (<1%), therefore further work is required to understand the remaining sources of phenotypic variance. There is also evidence of a “healthy volunteer” bias in UKB ^48^, especially among people who completed the mental health questionnaire ^31^, which may make this study not fully representative of the general population. Finally, the present study was focused on individuals of European ancestry therefore these results might not be valid for other ancestries^49^.

Despite the limitations, we report for the first time that smoking is associated with occurrence of psychotic experiences after adjusting for genetic risk for mental health, and that current smokers with higher genetic risk for depression and ADHD are more likely to experience delusions but not hallucinations. These results emphasise the importance of assessing environmental and genetic factors jointly and separately, and exploring particular types of psychotic experiences, as they can be differentially associated with smoking or other risk factors. This study also highlights the relationship between smoking and depression, and between smoking and ADHD on the occurrence of specific types of psychotic experiences in the general population, encouraging further studies to explore the effect of genetic risk for other psychiatric disorders on psychosis. Taken together, our results support the distinction of types of psychotic symptoms, the inclusion of environmental factors, and the study of cross-disorder genetic predictors to inform biological mechanisms underlying psychotic experiences.

## Data Availability

All data referred to in this manuscript is available upon request

https://github.com/juditperala/PRSxSmoking.

## Acknowledgments

This paper is dedicated to the memory of our dear friend and colleague Rob Keers, who sadly passed away while this manuscript was being peer-reviewed.We thank the UK Biobank participants who donated their time, experiences and DNA to this research, and to the UK Biobank team for making the data available. JGG would like to thank Dr Jonathan Coleman, Dr Christopher Hübel and Olakunle Oginni for valuable feedback on the analyses and the manuscript draft.

## Funding

JGG acknowledges support from the Queen Mary Principal’s Research Studentship in the School of Biological and Chemical Sciences. DMH is supported by a Sir Henry Wellcome Postdoctoral Fellowship (Reference 213674/Z/18/Z) and a 2018 NARSAD Young Investigator Grant from the Brain & Behavior Research Foundation (Ref: 27404). PBM acknowledges support from the National Institutes of Health Research (NIHR) Cardiovascular Biomedical Centre at Barts and The London, Queen Mary University of London (QMUL). JR acknowledges support from the European Union’s Horizon 2020 research and innovation programme under the Marie Sklodowska-Curie grant agreement No 786833. CHB acknowledges support from NIH grant No UO1 DA04440001A1. CHB is a member of the Royal Society Industry Fellows’ College. RK is supported by Wellcome Trust Grant 208881/Z/17/Z.

## Conflict of interests

The authors of this manuscript certify that they have NO affiliations with or involvement in any organization or entity with any financial interest.

## Notes

### Competing Interest Statement

The authors have declared no competing interest.

### Funding Statement

JGG is supported by a Queen Mary Principal's Research Studentship in the School of Biological and Chemical Sciences. DMH is supported by a Sir Henry Wellcome Postdoctoral Fellowship (Reference 213674/Z/18/Z) and a 2018 NARSAD Young Investigator Grant from the Brain & Behavior Research Foundation (Ref: 27404). PBM wishes to acknowledge support from the National Institutes of Health Research (NIHR) Cardiovascular Biomedical Centre at Barts and The London, Queen Mary University of London (QMUL). JR acknowledges support from the European Union's Horizon 2020 research and innovation programme under the Marie Sklodowska-Curie grant agreement No 786833. CHB acknowledges support from NIH grant No UO1 DA04440001A1. CHB is a member of the Royal Society Industry Fellows' College. RK is supported by Wellcome Trust Grant 208881/Z/17/Z.

### Summary of Updates

Summary of changes included in revised manuscript: - Main and interaction analyses now include PRSs for bipolar disorder (Stahl et al. 2019). - The different diagnoses included as psychotic disorders (schizophrenia, mania, hypomania, bipolar or manic depression, or any other type of psychotic disorder including schizotypal and delusional disorders) are now specified on first usage in the text. - A shortened version of the paragraphs that summarize the analysis pipeline has been moved to the introduction. - A new paragraph including Psychotic Experience ∼ PRS analyses has been included in the main effects section, and the prediction of psychotic experiences in UK Biobank using PRSs for schizophrenia, depression, ADHD and bipolar disorder were moved from Supplementary Figure 1 to Figure 2 in the main text. - Treatment with antidepressants was taken into account in the original submitted manuscript, since the presence of antidepressant drugs in the 'treatment medication' field was part of the inclusion criteria for depression (Supplementary Table 4). Depression was included as covariate in the association analyses between psychotic experiences and smoking behaviour. However, presence of antipsychotics in the treatment medication field was not part of the inclusion criteria for psychotic disorders. This has been amended now and antipsychotic treatment is included in the criteria for psychotic disorders (Supplementary Table 4). The list of antipsychotic drugs included is specified in Supplementary Table 5. Using the new criteria to ascertain cases for psychotic disorders, the number of cases increased from 4,847 to 6,727, but the study results remained similar. - We have checked the sample overlap between the depression GWAS and UK Biobank and assessed its impact. Sample overlap was 0.079% for the discovery sample (351/446,238), and 0.242% for the target sample (351/144,818). This has been included in the manuscript.

